# Improving The Care of Critically Ill Patients: Lessons Learned from The Promotion of Essential Emergency and Critical Care In Tanzania: A Qualitative Study

**DOI:** 10.1101/2024.05.24.24307887

**Authors:** Aneth Charles Kaliza, Linda Mlunde, Carl Otto Schell, Karima Khalid, Hendry Sawe, Elibariki Mkumbo Ba, Andrew Kigombola, Isihaka Mwandalima, Erasto Sylvanus, Said Kilindimo, Edwin Lugazia, Janeth Stanslaus Masuma, Tim Baker

## Abstract

**Objective:** To describe the lessons learned during the promotion of a new approach to the care of critically ill patients in Tanzania - Essential Emergency and Critical Care (EECC)

**Design:** A descriptive qualitative study using thematic analysis of structured interviews

**Setting and Participants:** The study was conducted in Tanzania, involving eleven policy makers, researchers and senior clinicians who participated in the promotion of EECC in the country.

**Results:** The five thematic lessons that emerged from the promotion of EECC in Tanzania were: (i) ensure early and close collaboration with the government and stakeholders; (ii) conduct research and utilize evidence; (iii) prioritize advocacy and address misconceptions about EECC; (iv) leverage events and embed activities in other health system interventions and (v) employ a multifaceted implementation strategy.

**Conclusion:** The results from this study show the efficacy of a holistic, comprehensive approach in promoting EECC as each strategy reinforces the others. This approach led the to the successful promotion of EECC and the development of a National Strategic Plan for EECC by the government of Tanzania.

**Article Summary:** Strengths and Limitations of this study:

*Strengths:*

- High credibility of findings due to the in-depth qualitative data collection process and the inclusion of diverse participants, which continued until data saturation was reached.
- Mitigation of personal biases by iterative sharing of findings with participants and key stakeholders

*Weaknesses:*

- The purposeful selection of participants may have missed some stakeholders with alternative viewpoints and experiences.
- We were unable to transcribe the interviews, instead, a codebook and audio recordings were used for cross-referencing which may have led some relevant information being missed.

## INTRODUCTION

Illness can progress in severity [1] if appropriate care is not provided. Progression to the most severe stage of any acute illness, regardless of a patient’s age, gender, social status, or medical condition is termed ‘critical illness’ [2]. Critical illness is characterized by vital organ dysfunction, a high risk of imminent death if care is not provided, and the potential for reversibility [3]. Critical illness is strikingly common – approximately one in ten patients either arriving to hospital or being treated in hospital are critically ill [4-6]. Globally, it is estimated that 45 million adults become critically ill each year [7]. While the burden of pediatric critical illness is unknown [8], sepsis and pneumonia are the leading causes of children’s mortality and seven million children require oxygen for pneumonia each year [9, 10]. The need for improving the care of critical illness globally and especially in low-resource settings is increasingly recognized [11-13].

To systematically improve care for critically ill patients, a focus has recently been proposed on Essential Emergency and Critical Care (EECC) – the care that should be provided to all critically ill patients in all hospitals in the world [1]. The EECC approach is guided by three principles: i) priority to those with the most urgent clinical need; ii) provision of life-saving treatments that support and stabilize failing vital organ functions when indicated; and iii) care to focus on effective actions of low cost and low complexity [1]. The COVID-19 pandemic, despite its devastating consequences, was a valuable source of learning for health systems as weaknesses were unveiled in the existing critical care model and the need for a more sustainable, inclusive and equitable critical care system [14, 15]. There are substantial gaps between the needs of critically ill patients and the availability of fundamental critical care in health systems in low and middle-income countries (LMICs) [16, 17]. EECC is an approach that could close these gaps, by focusing the human and other resources in healthcare towards the patients with the greatest clinical need – in line with a key goal for health systems of prioritizing resources to maximize health benefit [18]. The low costs of the 40 simple clinical processes contained in EECC, when compared to advanced critical care such as intensive care, enable care provision to many, even when resources are limited, and suggest that prioritizing EECC would be an equitable and cost-effective approach [19]. EECC is also important in settings of higher resources – to ensure that specialized medical services don’t overlook foundational care [2]. The concept of EECC has the potential to improve critical care in hospitals all over the world and substantially reduce preventable mortality [2].

In recent years, the Tanzanian government has been building a policy emphasis on the care of critically ill patients, with increased momentum due to the COVID-19 pandemic. Researchers have responded with the generation of evidence around critical illness and critical care in Tanzania [20-22] and the Tanzanian Ministry of Health has developed a National Strategic Plan for EECC Services [23]. The experiences of the introduction and promotion of EECC in Tanzania have provided insights into the challenges, opportunities, and potential solutions when introducing and promoting a new perspective to care, EECC. In this paper, we aim to systematically collect and analyze these experiences to determine the lessons learned for successful promotion of EECC, as a guide for researchers, policymakers, healthcare providers, and organizations involved in improving the care of critically ill patients worldwide.

## METHODS

### Study design and setting

We employed a qualitative descriptive design, conducting in-depth interviews with key informants in Tanzania from February to May 2023. Tanzania is a lower-middle income country in East Africa with a population of 65 million people, [24] a life expectancy of 67 years and an infant mortality rate of 33 deaths per 1000 live births. The country is a multi-party democratic republic with a long history of political stability.

### Study participants

Participants were purposefully selected based on their known contribution to the work around EECC in Tanzania. They included national-level researchers, healthcare professionals, policymakers, and representatives from implementing and development partners. All individuals who were approached participated in the study. During interviews, some participants suggested additional key informants involved in the promotion of EECC. The sample size was determined by the principle of saturation, when additional interviews were no longer expected to lead to new data.

### Data collection

Interviews were conducted in both English and Swahili, guided by a semi structured interview guide (Appendix). The guide encompassed questions pertaining to the strategies employed, stakeholders involved, facilitators and challenges encountered by participants during the promotion of EECC. The interview guide was piloted and adjusted before use.

The interviews took place either at the participants’ workplaces or via online calls for those inaccessible due to logistical constraints. Each interview lasted between 45 and 60 minutes. Prior to each interview, verbal consent was obtained from the participants, and interviews were audio recorded for accuracy and analysis purposes. The interview sessions were led by two researchers (AK and LM), both possessing backgrounds in clinical medicine and qualitative research methodologies.

### Data analysis

Following data collection, AK and LM immersed themselves in the interview data to enhance familiarity with the content by repeatedly listening to the audio recordings. Transcription was not possible due to logistical constraints. Analysis was conducted utilizing a rapid qualitative analysis approach. A deductive approach was employed to develop a codebook based on the interview guide questions. Researchers listened to the audio recordings and manually organized the content of the interview data into predefined codes derived from the codebook. A thematic approach was used to identify themes about the lessons learned. The emerging themes were discussed among the researchers and then shared with participants for their feedback to ensure their accuracy and resonance with their experiences.

## Ethical Considerations

Participants were provided with information about the study’s purpose procedures, and the interview guide prior to the interview. Verbal consent was obtained from each participant, ensuring they clearly understood the study, freely chose to participate, and they had an opportunity to ask questions and clarify any concerns before providing their consent. All collected data was securely stored and was only accessible to AK and LM. All personal identifiers were removed, and data were anonymized before analysis. The study was granted ethical approval from the Tanzanian National Institute for Medical Research (NIMR/HQ/R.8a/Vol.IX/3537).

## RESULTS

Eleven key informants were included, including researchers, implementation and development partners, policymakers, and emergency and critical care healthcare professionals (Table 1). Several participants had more than one area of expertise.

**Table 1:** Participant characteristics. Note the percentage adds up to more than 100% as several participants have more than one profession.

| Characteristics | Frequency n (%) N=11 |
| --- | --- |
| Female | 3 (27) |
| Profession |  |
| Researchers | 4 (36) |
| Emergency Physicians | 2 (18) |
| Critical Care Physicians | 4 (36) |
| Policy Makers | 6 (54) |
| Implementation partners | 4 (36) |

Five themes emerged from the data as lessons learned from the promotion of EECC in Tanzania: (1) ensure early and close collaboration with the government and stakeholders; (2) conduct research and utilize evidence; (3) prioritize advocacy and address misconceptions about EECC; (4) leverage events and embed activities in other health system interventions; and (5) use a multifaceted implementation strategy.

### Theme 1: Early and close collaboration with the government and stakeholders

Participants stressed the importance of early and continuous policy contact, fostering government leadership, plus contact with in-country stakeholders, United Nations agencies, and other partners. The EECC agenda was repeatedly communicated to policy makers and other stakeholders to maintain focus and conducting periodic dissemination meetings kept them informed of their research agenda and ongoing activities for the promotion of EECC.

*“… we did dissemination to get more people to understand the idea better. We invited people from [Universities], [government ministries], health facilities included in the study and other organizations. We introduced EECC to them, and they were very keen to make sure EECC is working” [participant no 1]*

For any country, the government has many agendas and areas that they are working on improving. To maintain focus on EECC, participants expressed that having an EECC expert based in-country was necessary for continuous contact with the government and stakeholders.

*“…it is important to establish early and continuous policy contact, for instance for the case of Tanzania, continuous contact about EECC with the Ministry of Health*.*” [participant no 5]*

Additionally, establishing and maintaining close contacts with partner organizations such as the United Nations Children’s Fund (UNICEF) helped to mobilize resources needed for promotion of EECC and the development of a national strategic plan for EECC.

*“… [funding] to the government in Zanzibar to do the first phase to devise the plan on how Zanzibar would increase EECC and also a lot of support and engagement with the government led to the National Strategic Plan for EECC*…*” [participant no 5]*

### Theme 2: Conduct research and utilize evidence

Participants emphasized the importance of conducting research to generate knowledge as a key strategy for promoting EECC. Researchers in Tanzania conducted baseline surveys to understand the gaps in the provision of critical care in Tanzania and neighboring countries; specifically mentioned was research in the provision of oxygen and availability of resources in facilities for the provision of EECC. The findings were disseminated to inform stakeholders about the need for EECC. Furthermore, health economics research to describe the potential cost and cost-effectiveness of EECC was found to be of particular interest to policymakers.

*“…we conducted baseline surveys to understand the status of critical care such as the critical care in low-income countries study done in 2009 which showed a lack the basic care and equipment for patients in hospitals*…*” [Participant no 5]*

*“…what we found is mostly what is needed might be present in the facility, but is not at the right place, and maybe not in the right amount*.*…” [participant no 1] “…research is not confined to knowledge generation, so it is important to develop an interface between research and policy, research and impact……ultimate job of a researcher is not just to create new knowledge, but to make the world a better place…” [participant no 5]*

*“…We have background information regarding the usefulness of EECC, which I think is one step good, and some conversation with the government regarding implementation of this which has already started*…*” [participant no 6]*

### Theme 3: Prioritize advocacy and address misconceptions about EECC

Advocacy emerged as a prominent theme during interviews with the informants. Participants highlighted that while working to influence decisions, policies, and practices in favor of EECC, there were four areas to address to avoid misconceptions:

I. The significance of clearly communicating what EECC is and ensuring its principles are well-understood. For instance, despite being a novel approach to critical care, the set of actions and treatments included in EECC are known practices in caring for sick patients. Additionally, it was found important to communicate that there are gaps in provision of EECC in both low and high-resource settings and that EECC is feasible in both settings and can be administered by any healthcare provider, regardless of their location within the hospital. *“*…*It’s a systems innovation, explaining the problem in a different way than it has been done before, providing a solution in a different way than it has been done before. It should be advocated everywhere in the world*…*” [participant no 5]*
II. Given that EECC embodies the most foundational processes in provision of care, there is a risk of it being overlooked and undervalued. Participants emphasized the need for clarity in this area while advocating for EECC. *“…There was this notion that this [EECC)] is already done, that it is not something that you should be talking about, people were dismissing it. They were like, we know that it is part of what we teach every day…But from experience, from what we have been looking into, from the study, we know that this is not happening all the time*…*” [participant no 8]*
III. The provision of EECC entails the reallocation of resources already present in the majority of hospitals, rather than necessitating additional resources. *“…A study was done that showed that this basic equipment is present in the hospitals but is either locked away or the person in charge is not around so they cannot be accessed*…*” [Participant no 7]* *“…we can make a difference in the outcome of our patients, we do not need to be rich to do that we have all the resources to make the difference, and EECC is trying to prove that…” [Participant no 9]*
IV. The terms “critical care” and “intensive care” are frequently used interchangeably in the health system, and it was not widely understood that critical care can be provided at a low cost anywhere in the hospital, rather than just in Intensive Care Units (ICUs). *“…for critical care, people understood that only particular staff should provide it, those in ICU and the emergency department. But even when you work in the outpatient department you should be able to identify a critically ill patient and manage him/her. They thought that when you have a critically ill patient you must send him/her to ICU…” [participant no 11]* *“…every time we are trying to get our message across it’s actually two messages. We must focus on critical illness, and we have to focus on the basics*… *if the world only hears the first one, that’s a challenge*…*” [Participant no 5]* *“…people should understand that even if you work in the ward or outpatient department, sometimes you can come across a critically ill patient, and you should be able to initiate something before even those who you regard can take care of that patient take over the management of that patient…” [Participant no 8]*

### Theme 4: Leverage events and embedded activities in other health system interventions

Participants explained how the increased global attention on emergency and critical care after the COVID-19 pandemic was harnessed to promote EECC. Additionally, they leveraged the focus in international health direction by the WHO for health emergency preparedness, response, and resilience during and after the pandemic.

*“…COVID-19 forced us to look at critical care with our own two eyes. After recognizing that patients need oxygen and we do not have oxygen, and for one to provide oxygen one needs nasal prongs, masks, and gas tanks which were not there, we even realized that we do not have good production of oxygen…” [participant no 10]*

Participants pointed out that they actively engaged with various technical working groups initiated by the Tanzania Ministry of Health in the COVID-19 response and had the opportunity to describe how EECC could enhance the emergency preparedness of health systems.

*“*…*we contacted people in the Emergency Preparedness committee to give our thoughts on the drafted guidelines on the treatment of COVID-19…” [participant no 5]*

*“…were invited to the Technical Working Groups during COVID-19 time, when they were preparing COVID-19 treatment guidelines… we started advocating for EECC telling them there would be many critically ill patients who would not benefit from advanced care…” [participant no 1]*

Participants explained that it was crucial that EECC was framed as a stepping-stone towards delivering optimal care and that it complemented already established processes in the health system and initiatives for improving care such as emergency obstetric care, Emergency Triage and Treatment (ETAT) of children, scaling up of emergency and intensive care units, and scale-up of oxygen ecosystems; facilitating such initiatives to be more effective and impactful.

*“*…*for example, when someone holds a training on malaria and they are explaining when we have a sick child with malaria, we must give antimalarial drugs, but we must also give EECC*…*” [Participant no 5]*

### Theme 5: Use a multifaceted implementation strategy

Participants said that employing a multifaceted approach is important when introducing a new concept like EECC. They felt that single interventions, (such as a training course, or the procurement of some equipment), are unlikely to have an impact or lead to sustainable change. They further noted that this approach enabled them to shape policy, contributing to the development of Tanzania’s ministry of health’s national strategic plan for EECC.

*“*… *to change policies and strategies, you need a countrywide, system effort to change and implement, from the top, Ministry, to regional local administration, to hospital administrators and providers, all at once…” [participant no 10]*

*“It is hard to make a system change, it needs a lot of processes, it needs patience, a lot of steps, advocacy, stakeholders buy-in, you have to try to find different ways and approaches to reach to your goals, it does not happen in a short time” [participant no 11]*

## DISCUSSION

We have found that a comprehensive approach including a multidisciplinary collaboration among a diverse group of professionals, policymakers, and implementation partners alongside employing multiple strategies, was able to successfully introduce and promote EECC in Tanzania. This approach ensured that interventions synergistically reinforced the others’ impacts and eventually led to the assimilation of the concept, the development of a National Strategic Plan for EECC Services and the increase in activities around EECC in the country. Such strategies that combine efforts to address various barriers have been seen to be more effective than interventions targeting one obstacle at a time [25], but can demand additional resources and effort [25]. Reducing complexity and tailoring approaches to address specific barriers, as done in Tanzania, has been recommended as a core principle of implementation science[25, 26].

Capitalizing on the global emphasis on critical care in the COVID-19 pandemic offered a unique opportunity to promote EECC. Despite the efforts to improve ICUs during the pandemic, most of these units faced major shortages of human resources and medical devices, jeopardizing both patients and healthcare workers’ safety [27]. Viewed as a stress test, the pandemic served as an opportunity to design more effective interventions to maximize scarce healthcare resources [28]. Based on this, an important research priority has been to develop feasible ways to identify early those who are critically ill, and those at risk of mortality [28]. The core aspects of EECC are the identification and care of the critically ill, and the provision of life-saving supportive care that is of low cost and low complexity [1].

When promoting EECC, misconceptions need to be addressed. The novelty of the EECC concept, with its cross-cutting prioritization of illness severity across all diagnoses, its emphasis on the most foundational care and its system-lens, can be at-odds with traditional approaches in health systems, leading to significant misunderstandings. EECC is care for critical illness, but it is not ICU care. The clinical processes for the provision of EECC are already familiar to healthcare professionals, potentially leading to confusion about its distinctiveness compared to existing approaches. The absence of a universally agreed-upon definition of critical care further amplifies misconceptions about EECC and its relationship to intensive care. The Tanzanian team, in their advocacy endeavors, placed a strong emphasis on explaining what EECC entails and what it does not, recognizing this as the foundation for dispelling misconceptions. The EECC approach prioritizes life-saving supportive care across all medical specialties, aiming to complement specialty-based care and bridge the quality gap between current practices and best-practice guidelines [1]. It is important to clarify that EECC is not advanced care, does not encompass the definitive care of underlying diagnoses, or intend to encompass all aspects of patient care.

### Strengths and Limitations

A strength of this study lies in the methodology of data collection which involved qualitative in depth until saturation was achieved during interviews. This method facilitated a profound understanding of the lessons learned, thereby enhancing credibility of the findings. Personal biases to data interpretation are common in research and we tried to mitigate this risk by iteratively sharing the evolving findings with study participants and stakeholders for their feedback to ensure that the reported results align with their lived experiences. The professional diversity of the study participants also yielded a wealth of material indicative of diverse perspectives. Our study has several limitations. First, the purposeful selection of participants may have resulted in a sample with similar perspectives on EECC, limiting the diversity of viewpoints and experiences that could be explored. Second, as participants were reflecting on their own experiences, there is a risk of subjectivity, which can lead to a one-sided view of the lessons under study. Participants may have also altered their reflections to align with what they perceived to be desirable to the interviewer. Some information may have been missed as interview transcriptions were not done.

## CONCLUSION

The integration of EECC into Tanzania’s health policy is a result of a holistic, comprehensive approach during its promotion. This approach included research and utilization of evidence, advocacy, embedding EECC in ongoing health system interventions, and, significantly, early, and close collaboration with the government and other stakeholders. The lessons from the Tanzania’s experience can be applied globally to improve critical care systems, foster access to care and optimal outcomes for all critically ill patients.

## Data Availability

As the project lacks ethical approval for public data sharing, we are unable to provide access to the data used in this study.

## Acknowledgements

We are grateful to the POETIC MUHAS team for their support since the inception of this study.

## AUTHOR CONTRIBUTIONS

AK and LM were responsible for designing the study, acquiring, analyzing, and interpreting the data. AK prepared the first draft of the manuscript, refined it based on input from co-authors, and submitted the final version. TB conceptualized and designed the study, contributed to data gathering, supported data analysis, and participated in the revision of the manuscript. CS, KK, HS, EM, AKa, IM, ES, SK, EL, and JM contributed to the acquisition of data and the revision of the manuscript. All authors have read and approved the final manuscript.

### APPENDIX: INTERVIEW GUIDE

#### Muhimbili University of Health and Allied Sciences

##### Greetings!

We would like to welcome you to a discussion about Essential Emergency and Critical Care (EECC) in Tanzania. You have been identified as one of the key stakeholders who has worked on promotion of Essential Emergency and Critical Care in the country. We would like to hear your experience of the process, what has been achieved and how.

##### Questions

1. When did you first hear about EECC? Was it a new concept to you?
2. In your opinion, why should we advocate for EECC in Tanzania?
3. Were there pushing factors to introducing EECC in Tanzania? For example, External/Societal (e.g. media, advocacy groups/quality) Probe: what led you to start the efforts of introducing EECC at the time you started?
4. Which strategies have been used to introduce EECC in Tanzania? Probes: Which activities did you do? How was the process?
5. Whom did you first approach to try to introduce the concept of EECC in Tanzania? What was their response? Who were the other people you approached? How did they respond?
  - Did these people know about EECC prior?
  - Was EECC found to be acceptable, appropriate, feasible by these people? Please explain.
  - In your opinion, why do you think they were able to prioritize EECC than other health care needs?
  - Were they ready to adopt, implement, and sustain it? How?
6. Did you have to make partnerships and connections to introduce EECC in Tanzania? What/who were they?
7. What were the economic, environmental, political, technological conditions that enabled you to introduce EECC in Tanzania? Probes: Was the process funded? Were there any policies and laws that supported the process? Were there any other facilitators for the process?
8. Which challenges did you encounter in the process of introducing EECC in Tanzania? Probe: What did not go well?
9. What were the successes of your efforts? Probes: What went well?
10. Were there any unintended outcomes during the process of introducing EECC in Tanzania? What were they?
11. What have you learned in the process of promoting EECC in Tanzania?
12. In your opinion, what is the status of EECC? Probe: What is going on with regards to EECC now?
13. In your opinion, what are the next steps regarding improving coverage of EECC in Tanzania and beyond? Probe: What do you envision to see as the future of EECC in Tanzania?
14. If you were to participate in the introduction of EECC in Tanzania all over again, what would you have done differently?
15. What recommendations do you have for others who want to introduce the concept of EECC in their settings?
16. Who else did you work with to promote EECC in Tanzania?
17. Is there anything else that you would like to share with us regarding EECC in Tanzania?

Thank you so much for your time!

